# Outpatient Portal Use in Prenatal Care: Differential Use by Race, Risk, and Area Social Determinants of Health Factors

**DOI:** 10.1101/2021.05.25.21257657

**Authors:** Priti Singh, Pallavi Jonnalagadda, Evan Morgan, Naleef Fareed

## Abstract

**Objective:** To report the relationship of Outpatient Patient Portal (OPP) use with clinical risk, area social determinants of health (SDoH), and race/ethnicity among pregnant women.

**Methods:** Regression models predicting overall and individual portal feature use (main effects and interactions) based on key variables were specified using logfiles and clinical data.

**Results:** Overall OPP use among non-Hispanic Black women or patients who lived in lower SDoH neighborhoods were significantly less. High-risk pregnancy patients were likely to use the OPP more than those with normal-risk pregnancy. We found similar associations with individual OPP features, like the Visit (scheduling) and My Record (test results). We also found significant interactive associations between race, clinical risk, and SDoH. Non-Hispanic Black women living in lower SDoH areas used OPP less than non-Hispanic White women from similar or affluent areas.

**Conclusion:** More research must be conducted to learn of OPP implications for pregnant women with specific clinical diagnoses.

## Introduction

Pregnancy represents a critical period of care when active health engagement can influence maternal and infant outcomes. Patient portals promote improved patient-provider communication, shared decision making, and patient engagement [1]. Depending on the type of portal, patients can access medication lists, information about recent doctor visits, immunization history, as well as securely message one’s doctor, refill prescriptions, make payments, and schedule non-urgent appointments [2]. Patient portals provide patients with the means to learn about their health and participate in their health care. Higher portal use was demonstrated among pregnant women, who had access to personalized health information through the portal as compared to those who had general information [3]. Few studies examined patterns of portal use in obstetric (OB) patients or specific factors that can influence use among these patients [3–5].

Patient portal use has been associated with improved medication adherence, disease awareness, self-management of the disease, and patient satisfaction. Their use has been associated with decreased office visits and improved preventative care [6]. Although less studied, patient portals have also shown promise in the improvement of clinical outcomes. For instance, portal use was associated with improved glycemic control [7]. Studies have assessed outpatient portal (OPP) use by examining number of logins [8] and engagement metrics generated from log file analysis [9–12].

The digital divide — the gulf between people who have and do not have access to information technologies [13]— may exist among patient portal users and further exacerbate health inequities across communities. OPP use was less likely among racial/ethnic minorities and individuals living in high deprivation neighborhoods – a proxy for social determinants of health (SDoH) [14]. Residential racial segregation is particularly high for African-Americans, perpetuating differences in socioeconomic status (SES) and health disparities [15]. Use of patient portals is at the intersection of health status, race/ethnicity, and SDoH. Patient portals may be disproportionately used among specific groups of pregnant women with already better health status and greater likelihood of better health outcomes [16].

The primary objective of this work is to report on the relationship of OPP use with maternal clinical risk, SDoH (as measured by an area deprivation index) and racial/ethnic identity. We hypothesize that OPP use will be higher among women at greater risk of pregnancy-related complications, lower among women living in deprived neighborhoods, and vary by the race/ethnic identity of patients. We further explore the influence of the intersection of clinical risk, SDoH, and racial identity on OPP use at our Academic Medical Center (AMC). It is important to study how this novel health information technology is used among OB patients to identify areas of improvement to ensure that patient engagement with their health and health care is equitable.

## Methods

This study is a secondary analysis of electronic health records (EHR) and audit log data on My Chart portal usage for women who received prenatal care at our AMC. The log provides a temporal sequence, through the use of time stamps, of specific actions undertaken by patients on the portal. A retrospective cohort of 17,132 pregnant women seen by the Department of Obstetrics and Gynecology (OB/GYN) and Maternal Fetal Medicine (MFM) Division at our AMC, during the study period of 1/1/2016 to 8/1/2020 were analyzed for their OPP use. These patients had MyChart accounts through our AMC.

### Study population

The study population included information on women who had a) at least one episode with a birth at the AMC; b) visited the AMC OB/GYN or MFM for prenatal care at least once; c) were registered users of AMC MyChart portal; d) were at least 18 years of age. Outpatient visits for each patient are recorded using unique account identifiers, while Medical Record Numbers (MRNs) are used to link these visits to outpatient portal encounters. Portal activities for 20,788 unique pregnancy episodes from our AMC for were retrieved from our Information Warehouse, out of which complete information was available on 9,712 patient episodes. Demographic information of these study patients are included in Table 1.

**Table 1:** Outpatient portal features and their descriptions

| Portal Feature Group | Description |
| --- | --- |
| Messaging | Links to messaging center, letters to the patient, prescription refill option |
| Visits | List of past and upcoming visits |
| My Record | List of medications, allergies; medical history, immunizations; test results and health summary; preventive care and a summary of plan of care |
| Medical Tools | Share medical records with other services; participate in research studies; and connect tracking devices |
| Billing | Account summary, payment |
| Resources | Terms and conditions; patient education; and frequently asked questions |
| Proxy | Request or renew proxy |
| Preferences | Personal and security settings; notification preferences |
| Custom | Miscellaneous |

### My Chart Patient Portal

Our AMC’s MyChart (Epic Systems: Verona, Wisconsin) portal is a free, personalized, and secure electronic system that is tethered to a patient’s personal medical record. It is designed to facilitate patient-provider communication through features including electronic message exchange, appointment scheduling, availability of test results, billing, and more.

### Data processing

Our analytical data set contained details about encounter summaries of outpatient visit for patients with activated portal accounts. It included information on time and duration of portal use, features accessed, user agent information, test results, and demographic information. These data were processed using the data mining technique described by DiTosto and colleagues to examine patient engagement based on patient interaction with the portal tool [11]. Based on their prescribed methodology, the portal features were grouped into nine areas summarized in table 1.

### Data analysis

We use a negative binomial regression model specification to account for the overly dispersed portal use variables. The model is offset by weeks of pregnancy because portal use could vary among patients based on the duration of prenatal care. Our fully-adjusted models include episode of pregnancy, age at first clinical encounter, Charlson Comorbidity Index (CCI), a area SDoH rank based on ZIP code [17], pregnancy risk status (high/normal), race/ethnicity (non-Hispanic White, non-Hispanic Black, Hispanic and Others) and body mass index at the initial encounter.

The area SDoH rank is the area deprivation index (ADI) that represents rankings of neighborhoods within the AMC’s state. We converted the ADI to quantiles. The measure was retrieved from the Neighborhood Atlas [17]. High-risk pregnancy included women with multiple pregnancies, diabetes, high blood pressure, genetic conditions, history of premature birth, preeclampsia, advanced maternal age, or any condition requiring high-risk care or fetal treatment. We report the estimated incidence rate ratios (IRR) from fully-adjusted models along with corresponding 95% confidence intervals. We tested the interaction between race/ethnicity, risk status and ADI. We also performed a sensitivity analysis to see if our results differed due to the COVID-19 pandemic by excluding any pregnancies with births after January, 2020.

## Results

Demographic information for our sample is presented in Table 2. Based on our total analytical sample, 89% had normal-pregnancy related risks while 11% had high-pregnancy related risks. A majority of the women with activated portal accounts at the AMC were non-Hispanic Whites, followed by non-Hispanic Blacks and Hispanics. In almost 67% of the episodes, women visited their obstetrician within their first trimester for initiation of prenatal care. These patterns were consistent across the different risk groups.

**Table 2:** Descriptive Statistics for Study Samples

| **Patient Characteristics** | **Total**<br>**N=9,712** | **Normal Risk**<br>**N=8,624** | **High Risk**<br>**N=1,088** |
| --- | --- | --- | --- |
| **Age** (mean, SD) | 30 (5) | 30 (5) | 31 (5) |
| **Race/ethnicity** (n, %) |  |  |  |
| Non-Hispanic White | 6,590 (67.85%) | 5,873 (68.10%) | 717 (65.90%) |
| Non-Hispanic Black | 1,868 (19.23%) | 1,672 (19.39%) | 196 (18.01%) |
| Hispanic | 1,210 (12.46%) | 1,041 (12.07%) | 169 (15.53%) |
| Other | 44 (0.45%) | 38 (0.44%) | 6 (0.55%) |
| **Gestational age at initiation of care at AMC** (n, %) |  |  |  |
| 1<sup>st</sup> trimester | 6,436 (66.43%) | 5,757 (66.90%) | 679 (62.70%) |
| 2<sup>nd</sup> trimester | 2,530 (26.11%) | 2,186 (25.40%) | 344 (31.76%) |
| 3<sup>rd</sup> trimester | 723 (7.46%) | 663 (7.70%) | 60 (5.54%) |
| **Clinical risk (n, %)** |  |  |  |
| Normal | 8,624 (88.80%) | - | - |
| High | 1,088 (11.20%) | - | - |
| **Area SDoH Rank (n, %)** |  |  |  |
| Quantile 1 | 2,464 (25.37%) | 2,213 (25.66%) | 251 (23.07%) |
| Quantile 2 | 2,187 (22.52%) | 1,972 (22.87%) | 215 (19.76%) |
| Quantile 3 | 1,906 (19.63%) | 1,667 (19.33%) | 239 (21.97%) |
| Quantile 4 | 1,860 (19.15%) | 1,626 (18.85%) | 234 (21.51%) |
| Quantile 5 | 1,295 (13.33%) | 1,146 (13.39%) | 149 (13.69%) |

Note: High risk pregnancy includes women with either multiple pregnancies, diabetes, high blood pressure, genetic conditions, history of premature birth, preeclampsia, advanced maternal age, or any condition requiring high-risk care or fetal treatment. SDoH = Social Determinants of Health

Estimated IRRs for overall portal use and individual features are presented in Table 3. We found that non-Hispanic Black women used the portal (IRR: 0.74; 95% CI: 0.70, 0.78) less than non-Hispanic White women (Figure 1a). Women with high-risk pregnancies used the portal 46% more (IRR: 1.46; 95% CI:1.37-1.55) than those with normal pregnancy risks. Area SDoH conditions also played a significant role in portal use. Women living in the most deprived areas used the portal 22% less than those living in the least deprived ones (IRR 0.79: CI: 0.72-0.87).

**Table 3:**
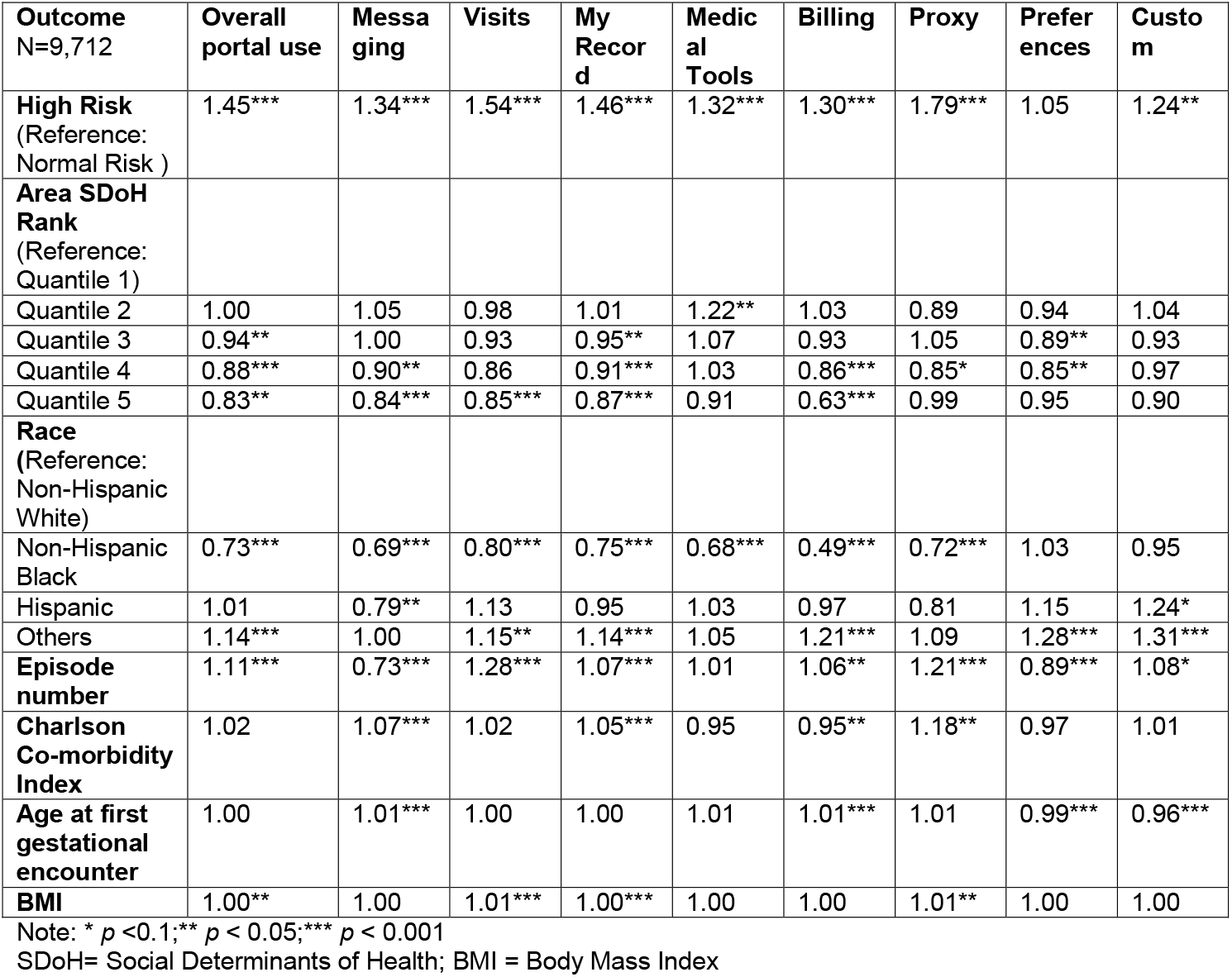
Rate ratios for outpatient portal use outcomes and key study variables

| **Outcome**<br>N=9,712 | **Overall**<br>**portal use** | **Messa**<br>**ging** | **Visits** | **My**<br>**Recor**<br>**d** | **Medic**<br>**al**<br>**Tools** | **Billing** | **Proxy** | **Prefer**<br>**ences** | **Custo**<br>**m** |
| --- | --- | --- | --- | --- | --- | --- | --- | --- | --- |
| **High Risk**<br>(Reference:<br>Normal Risk ) | 1.45*** | 1.34*** | 1.54*** | 1.46*** | 1.32*** | 1.30*** | 1.79*** | 1.05 | 1.24** |
| **Area SDoH**<br>**Rank**<br>(Reference:<br>Quantile 1) |  |  |  |  |  |  |  |  |  |
| Quantile 2 | 1.00 | 1.05 | 0.98 | 1.01 | 1.22** | 1.03 | 0.89 | 0.94 | 1.04 |
| Quantile 3 | 0.94** | 1.00 | 0.93 | 0.95** | 1.07 | 0.93 | 1.05 | 0.89** | 0.93 |
| Quantile 4 | 0.88*** | 0.90** | 0.86 | 0.91*** | 1.03 | 0.86*** | 0.85* | 0.85** | 0.97 |
| Quantile 5 | 0.83** | 0.84*** | 0.85*** | 0.87*** | 0.91 | 0.63*** | 0.99 | 0.95 | 0.90 |
| **Race**<br>(Reference:<br>Non-Hispanic<br>White) |  |  |  |  |  |  |  |  |  |
| Non-Hispanic<br>Black | 0.73*** | 0.69*** | 0.80*** | 0.75*** | 0.68*** | 0.49*** | 0.72*** | 1.03 | 0.95 |
| Hispanic | 1.01 | 0.79** | 1.13 | 0.95 | 1.03 | 0.97 | 0.81 | 1.15 | 1.24* |
| Others | 1.14*** | 1.00 | 1.15** | 1.14*** | 1.05 | 1.21*** | 1.09 | 1.28*** | 1.31*** |
| **Episode**<br>**number** | 1.11*** | 0.73*** | 1.28*** | 1.07*** | 1.01 | 1.06** | 1.21*** | 0.89*** | 1.08* |
| **Charlson**<br>**Co-morbidity**<br>**Index** | 1.02 | 1.07*** | 1.02 | 1.05*** | 0.95 | 0.95** | 1.18** | 0.97 | 1.01 |
| **Age at first**<br>**gestational**<br>**encounter** | 1.00 | 1.01*** | 1.00 | 1.00 | 1.01 | 1.01*** | 1.01 | 0.99*** | 0.96*** |
| **BMI** | 1.00** | 1.00 | 1.01*** | 1.00*** | 1.00 | 1.00 | 1.01** | 1.00 | 1.00 |

Note: \* $p < 0.1$ ; \*\* $p < 0.05$ ; \*\*\* $p < 0.001$
SDoH= Social Determinants of Health; BMI = Body Mass Index

**Figure 1.**
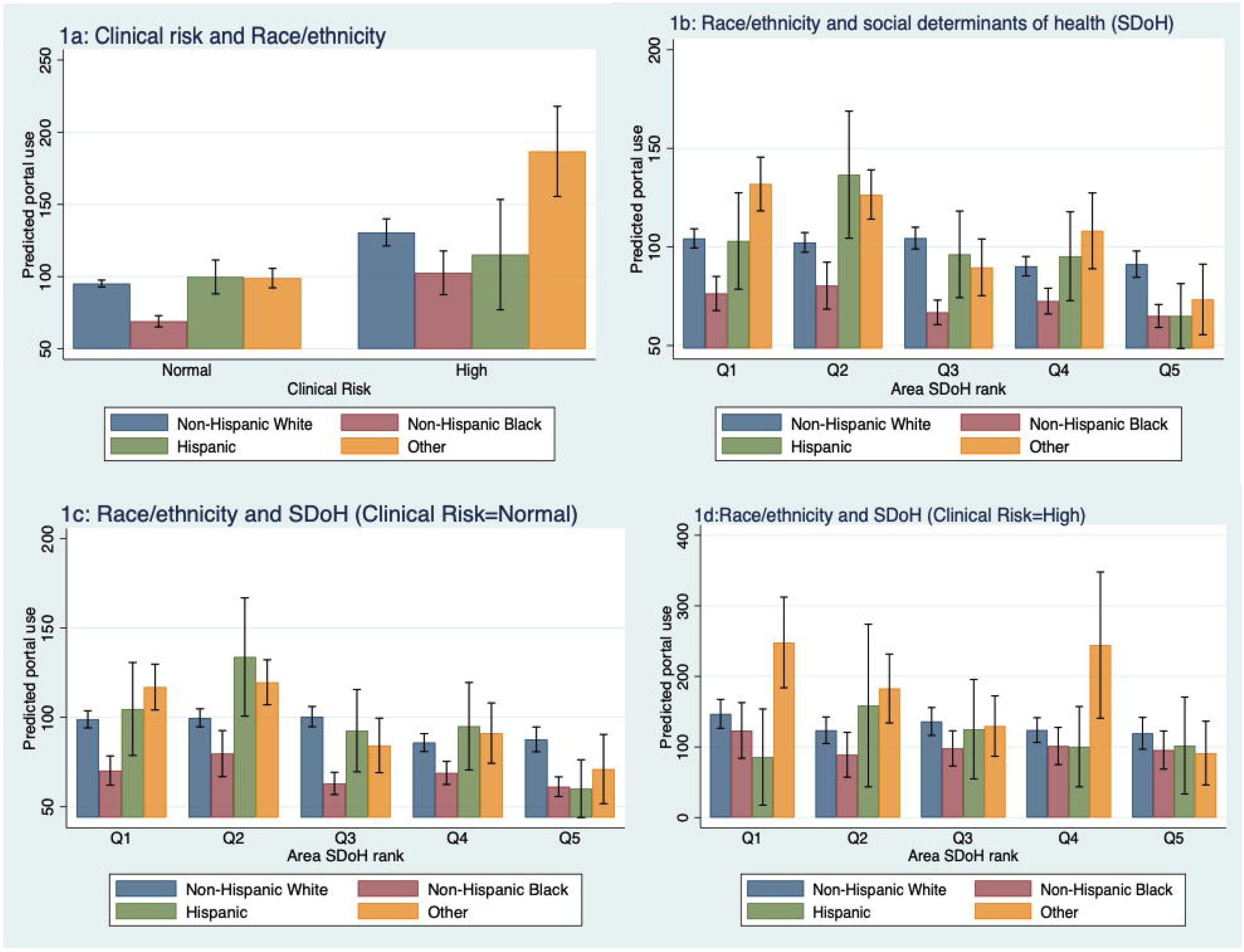
Interactive plots for overall outpatient portal use by race, area social determinants of health, and clinical risk for pregnant women. Panel 1a: Interaction plot for Risk and Race/ethnicity (1=Non-Hispanic White; 2=Non-Hispanic Black; 3=Hispanic; 4=Other); Panel 1b: Interaction plot for Race/ethnicity & Area SDoH rank (Q1: best and Q5: worst); Panel 1c: Interaction plot for Race/ethnicity & Area SDoH rank when clinical risk is normal; Panel 1d: Interaction plot for Race/ethnicity & Area SDoH rank when clinical risk is high

Based on counts of individual features, the Visit feature, which involved scheduling information, was most used, followed by My Record, that helped patients manage their medication and test results (results not shown). Similar to our results for overall portal use, we found that these top features were less used among non-Hispanic Black women in comparison to non-Hispanic White women. Women from the most deprived areas used these features less than women living in less deprived areas. We observed similar patterns for race for several other features. Women who had subsequent pregnancies had a lower use of the messaging feature, but a higher use of the visits feature. No activity was observed for the Resources feature across any patients.

Two and three-way interactions between race/ethnicity, clinical risk, and area SDoH were also examined. We found statistically significant interactions displayed in Figure 1 (see Table S1 for results from our interaction models). As risk of complications increased, pregnant women of the Other race group used the portal significantly more than non-Hispanic White women with similar risks (IRR: 1.44; 95% CI: 1.04-2.00), shown in Figure 1a. Women from the Other race group, who lived in least deprived areas used the portal more than non-Hispanic White women living in similar neighborhoods. However, with increased levels of area SDoH deprivation, portal use in women from Other race group reduced significantly (Figure 1b). Within the normal risk category, as deprivation increased, overall portal use was significantly lower among women of the Other race group compared to non-Hispanic White women (Figure 1c). No significant interactions were seen between race/ethnicity and ADI within the high-pregnancy related risk group (Figure 1d). Irrespective of risk and ADI, portal use was consistently lower among non-Hispanic Black women than other race groups. Similar patterns were observed for the billing feature; portal use was likely to reduce with deprivation (results not shown).

In regard to our sensitivity analysis, we found the pre-COVID estimates to be similar to those from our main analysis (see Table S2 for details).

## Discussion

We found non-Hispanic White women to be among the lowest portal users for pre-natal care and this pattern was consistent across different area SDoH and risk levels. Our results suggest that interaction of neighborhood and race impacts portal use. Despite increased pregnancy-related risks, non-Hispanic Black women used the portal less, even lower than non-Hispanic White women with high or normal pregnancy risk, a group that seemingly required less HIT and provider engagement. On the other hand, higher overall portal use among Other or Hispanic women suggests that not all women experienced the aforementioned phenomenon.

Prior studies of portal use in low-income populations found disparities by race and SES in OPP use, where higher use was associated with White race and being insured [18]. We found that area SDoH was associated with lower OPP use. Previous evidence suggests that better access to broadband internet [14] and smartphones with data, [19,20] can help bridge health disparities [21–23], including the use of patient portals. Qualitative research suggests that a one-time introduction to OPP is inadequate, especially among low-income pregnant women. Moreover, these women have also expressed a desire for consistent patient education and support that is tailored to their OB histories [24].

We observed greater OPP use among women at higher risk of pregnancy complications. Prior work suggests that use of personal health records was greater among those with chronic conditions [25]. Evidence from a study on portal use and glycemic control in prenatal care suggested that secure messaging is higher among those with suboptimal glucose control [5]. More research is needed to explore the implications of portal use for health outcomes among women with high-risk conditions and whether there are differential effects based on types of clinical diagnoses presented at pregnancy.

## Limitations

Our analysis was focused on OB patients in a single health system using an OPP from one particular vendor, thereby limiting the generalizability of our findings. Also, ADI is a proxy for SDoH factors and may not completely capture individual SDoH factors.

## Conclusion

OB patients interact with the healthcare system for nearly nine to ten months. HIT tools like patient portals can facilitate these interactions. Future studies should further explore how patient portal use based on race/ethnicity, area SDoH, and clinical risk can have differential effects on patient experiences and health outcomes.

## Data Availability

Data is restricted for this study.

## References

1 Wade-Vuturo AE, Mayberry LS, Osborn CY. Secure messaging and diabetes management: experiences and perspectives of patient portal users. J Am Med Inform Assoc 2013;20:519–25. doi:10.1136/amiajnl-2012-001253

2 What is a patient portal? | HealthIT.gov. https://www.healthit.gov/faq/what-patient-portal (accessed 26 Mar2021).

3 Shaw E, Howard M, Chan D, et al. Access to Web-Based Personalized Antenatal Health Records for Pregnant Women: A Randomized Controlled Trial. J Obstet Gynaecol Can 2008;30:38–43. doi:10.1016/S1701-2163(16)32711-6

4 Ukoha EP, Feinglass J, Yee LM. Disparities in electronic patient portal use in prenatal care: retrospective cohort study. J Med Internet Res 2019;21:e14445. doi:10.2196/14445

5 Holder K, Ukoha E, Feinglass J, et al. Relationship between patient portal utilization and glycemic control outcomes during pregnancy. J Diabetes Sci Technol 2021;:1932296821998742. doi:10.1177/1932296821998742

6 Kruse CS, Bolton K, Freriks G. The effect of patient portals on quality outcomes and its implications to meaningful use: a systematic review. J Med Internet Res 2015;17:e44. doi:10.2196/jmir.3171

7 Graetz I, Huang J, Muelly ER, et al. Association of mobile patient portal access with diabetes medication adherence and glycemic levels among adults with diabetes. JAMA Netw Open 2020;3:e1921429. doi:10.1001/jamanetworkopen.2019.21429

8 Lau M, Campbell H, Tang T, et al. Impact of patient use of an online patient portal on diabetes outcomes. Can J Diabetes 2014;38:17–21. doi:10.1016/j.jcjd.2013.10.005

9 Jones JB, Weiner JP, Shah NR, et al. The wired patient: patterns of electronic patient portal use among patients with cardiac disease or diabetes. J Med Internet Res 2015;17:e42. doi:10.2196/jmir.3157

10 Sieverink F, Kelders S, Poel M, et al. Opening the black box of electronic health: collecting, analyzing, and interpreting log data. JMIR Res Protoc 2017;6:e156. doi:10.2196/resprot.6452

11 Di Tosto G, McAlearney AS, Fareed N, et al. Metrics for outpatient portal use based on log file analysis: algorithm development. J Med Internet Res 2020;22:e16849. doi:10.2196/16849

12 Tsai R, Bell EJ, Woo H, et al. How patients use a patient portal: an institutional case study of demographics and usage patterns. Appl Clin Inform 2019;10:96–102. doi:10.1055/s-0038-1677528

13 van Dijk J, Hacker K. The digital divide as a complex and dynamic phenomenon. The Information Society 2003;19:315–26. doi:10.1080/01972240309487

14 Perzynski AT, Roach MJ, Shick S, et al. Patient portals and broadband internet inequality. J Am Med Inform Assoc 2017;24:927–32. doi:10.1093/jamia/ocx020

15 Williams DR, Collins C. Racial residential segregation: a fundamental cause of racial disparities in health. Public Health Rep 2001;116:404–16. doi:10.1093/phr/116.5.404

16 Brewer LC, Fortuna KL, Jones C, et al. Back to the future: achieving health equity through health informatics and digital health. JMIR Mhealth Uhealth 2020;8:e14512. doi:10.2196/14512

17 Kind AJH, Buckingham WR. Making Neighborhood-Disadvantage Metrics Accessible - The Neighborhood Atlas. N Engl J Med 2018;378:2456–8. doi:10.1056/NEJMp1802313

18 Ancker JS, Barrón Y, Rockoff ML, et al. Use of an electronic patient portal among disadvantaged populations. J Gen Intern Med 2011;26:1117–23. doi:10.1007/s11606-011-1749-y

19 Cell phone ownership hits 91% of adults | Pew Research Center. https://www.pewresearch.org/fact-tank/2013/06/06/cell-phone-ownership-hits-91-of-adults/ (accessed 23 Feb2021).

20 10% of Americans don’t use the internet | Pew Research Center. https://www.pewresearch.org/fact-tank/2019/04/22/some-americans-dont-use-the-internet-who-are-they/ (accessed 24 Jan2020).

21 Chou W-YS, Liu B, Post S, et al. Health-related Internet use among cancer survivors: data from the Health Information National Trends Survey, 2003-2008. J Cancer Surviv 2011;5:263–70. doi:10.1007/s11764-011-0179-5

22 Prestin A, Vieux SN, Chou W-YS. Is online health activity alive and well or flatlining? findings from 10 years of the health information national trends survey. J Health Commun 2015;20:790–8. doi:10.1080/10810730.2015.1018590

23 Fareed N, Swoboda CM, Jonnalagadda P, et al. Persistent digital divide in health-related internet use among cancer survivors: findings from the Health Information National Trends Survey, 2003-2018. J Cancer Surviv 2021;15:87–98. doi:10.1007/s11764-020-00913-8

24 Kim J, Mathews H, Cortright LM, et al. Factors Affecting Patient Portal Use Among Low-Income Pregnant Women: Mixed-Methods Pilot Study. JMIR Formativ Res 2018;2:e6. doi:10.2196/formative.5322

25 Archer N, Fevrier-Thomas U, Lokker C, et al. Personal health records: a scoping review. J Am Med Inform Assoc 2011;18:515–22. doi:10.1136/amiajnl-2011-000105

