## Supplementary material for "Outpatient Portal Use in Prenatal Care: Differential Use by Race, Risk, and Area Social Determinants of Health Factors": Table

**Supplementary Tables**

**Table S1: Regression outcomes with interaction effects**

| **Outcome** | **Overall portal use** | **Messaging** | **Visits** | **My Record** | **Medical Tools** | **Billing** | **Proxy** | **Preferences** | **Custom** |
| --- | --- | --- | --- | --- | --- | --- | --- | --- | --- |
| **Episode number** | 1.14*** | 0.73*** | 1.28** | 1.07*** | 1.02** | 1.06** | 1.23*** | 0.89** | 1.09** |
| **CCI** | 1.03 | 1.08*** | 1.01 | 1.05** | 0.96 | 0.95** | 1.20*** | 0.98 | 1.01 |
| **SDoH Rank** |  | | | | | | | | |
| Q1 | Reference | | | | | | | | |
| Q2 | 1.01 | 1.04 | 1.00 | 1.01 | 1.23** | 1.00 | 0.88 | 1.00 | 1.11 |
| Q3 | 1.01 | 1.04 | 1.02 | 0.98 | 1.09 | 1.00 | 1.18 | 0.92 | 1.00 |
| Q4 | 0.87*** | 0.94 | 0.83*** | 0.89** | 1.03 | 0.86** | 0.77** | 0.94 | 0.96 |
| Q5 | 0.88** | 0.83** | 0.93 | 0.92* | 1.03 | 0.70*** | 0.95 | 1.07 | 1.00 |
| **Race/ethnicity** |  | | | | | | | | |
| Non-Hispanic White | Reference | | | | | | | | |
| Non-Hispanic Black | 0.71*** | 0.57*** | 0.78** | 0.71*** | 0.62** | 0.51*** | 1.04 | 1.0 | 1.12 |
| Hispanic | 1.06 | 0.86 | 1.25 | 0.91 | 1.09 | 0.975 | 1.10 | 1.46 | 1.29 |
| Other | 1.18** | 1.09 | 1.16* | 1.16** | 1.16 | 1.35*** | 0.71* | 1.59*** | 1.31** |
| **SDoH Rank*Race/ethnicity** |  | | | | | | | |  |
| Q2* Non-Hispanic Black | 1.12 | 1.42** | 1.12 | 1.13 | 0.96 | 1.13 | 0.71 | 1.24 | 0.71 |
| Q2* Hispanic | 1.27 | 1.19 | 1.19 | 1.45** | 1.73 | 1.31 | 1.00 | 0.68 | 1.05 |
| Q2* Other | 1.01 | 0.98 | 1.08 | 0.97 | 0.94 | 1.00 | 1.88** | 0.71** | 0.85 |
| Q3* Non-Hispanic Black | 0.89 | 1.14 | 0.82 | 0.96 | 1.23 | 0.77** | 0.45** | 0.98 | 0.70* |
| Q3* Hispanic | 0.87 | 1.06 | 0.78 | 0.97 | 0.64 | 0.95 | 0.30** | 0.66 | 0.88 |
| Q3* Other | 0.71** | 0.63** | 0.72** | 0.83 | 0.80 | 0.54*** | 0.72 | 0.82 | 0.86 |
| Q4* Non-Hispanic Black | 1.13 | 1.11 | 1.19 | 1.13 | 1.22 | 1.04 | 0.76 | 0.91 | 1.04 |
| Q4* Hispanic | 1.04 | 0.69 | 1.02 | 1.16 | 0.81 | 1.14 | 0.53 | 0.61 | 0.82 |
| Q4* Other | 0.90 | 0.90 | 0.95 | 0.94 | 0.75 | 0.64** | 2.28** | 0.62** | 1.10 |
| Q5* Non-Hispanic Black | 0.98 | 1.29** | 0.93 | 1.01 | 0.92 | 0.81* | 0.60** | 0.89 | 0.75 |
| Q5* Hispanic | 0.65** | 0.76 | 0.61** | 0.71 | 0.58 | 0.54** | 0.45 | 0.74 | 0.59 |
| Q5* Other | 0.68** | 0.66** | 0.66** | 0.72** | 0.46 | 0.69* | 0.62 | 0.45** | 0.79 |
| **Risk** |  | | | | | | | | |
| Normal | Reference | | | | | | | | |
| High | 1.47*** | 1.36 | 1.66*** | 1.44*** | 1.25 | 1.17 | 0.80 | 1.00 | 1.39* |
| **SDoH Rank*Risk** |  |  |  |  |  |  |  |  |  |
| Q2*High Risk | 0.83 | 1.03 | 0.72** | 0.87 | 0.96 | 1.07 | 0.91 | 1.00 | 0.59** |
| Q3*High Risk | 0.92 | 0.97 | 0.88 | 0.98 | 1.10 | 0.93 | 1.48 | 1.18 | 0.96 |
| Q4*High Risk | 0.99 | 0.88 | 0.97 | 1.03 | 0.98 | 1.03 | 2.39** | 0.98 | 0.72 |
| Q5*High Risk | 0.93 | 0.90 | 0.85 | 0.95 | 1.23 | 1.01 | 4.94*** | 0.81 | 0.69 |
| **Race/ethnicity*Risk** |  | | | | | | | | |
| Non-Hispanic Black*High Risk | 1.22 | 1.59* | 1.23 | 1.16 | 2.21 | 1.09 | 0.69 | 1.22 | 0.40* |
| Hispanic *High Risk | 0.58 | 0.48 | 0.41 | 0.94 | 0.88 | 0.49 | 0.47 | 1.72 | 0.58 |
| Other*High Risk | 1.44** | 1.35 | 1.36 | 1.51** | 1.15 | 1.35 | 7.3*** | 1.20 | 1.18 |
| **SDoH Rank*Race/ethnicity*Risk** |  | | | | | | | | |
| Q2* Non-Hispanic Black*High Risk | 0.73 | 0.43** | 0.76 | 0.73 | 0.08** | 0.96 | 0.88 | 0.16** | 2.07 |
| Q2*Hispanic *High Risk | 1.67 | 0.83 | 2.38 | 0.99 | 0.32 | 2.14 | 22.78 | 1.75 | 7.79 |
| Q2*Other*High Risk | 0.86 | 0.51** | 1.00 | 0.95 | 1.00 | 0.78 | 0.13** | 1.09 | 2.28 |
| Q3* Non-Hispanic Black*High Risk | 0.93 | 0.67 | 0.87 | 0.99 | 0.37 | 1.38 | 2.27 | 0.99 | 2.82 * |
| Q3*Hispanic *High Risk | 1.75 | 1.35 | 2.79 | 1.00 | 0.57 | 1.81 | 6.99 | 0.90 | 2.24 |
| Q3*Other*High Risk | 0.80 | 0.99 | 0.68 | 0.68 | 1.05 | 1.32 | 1.23 | 0.67 | 0.66 |
| Q4* Non-Hispanic Black*High Risk | 0.84 | 0.72 | 0.79 | 0.80 | 0.51 | 1.09 | 2.66 | 0.75 | 2.51 |
| Q4*Hispanic *High Risk | 1.24 | 2.13 | 1.51 | 0.80 | 2.22 | 1.90 | 0.76 | 0.32 | 2.78 |
| Q4*Other*High Risk | 1.28 | 0.90 | 1.38 | 1.09 | 2.13 | 1.98 | 0.04** | 1.08 | 2.51 |
| Q5* Non-Hispanic Black*High Risk | 0.94 | 0.96 | 0.96 | 0.83 | 0.30 | 1.61 | 0.93 | 0.79 | 3.71** |
| Q5*Hispanic *High Risk | 2.18 | 1.93 | 3.03 | 1.32 | 1.36 | 2.72 | 3.56 | 0.96 | 6.74 |
| Q5*Other*High Risk | 0.66 | 0.59 | 0.67 | 0.74 | 0.34 | 0.70 | 0.6** | 2.07 | 1.88 |
| **Age at First Encounter** | 1.00 | 1.02*** | 1.00 | 1.00 | 1.01 | 1.02*** | 1.00 | 0.99*** | 0.96*** |
| **BMI** | 1.00** | 1.00 | 1.01*** | 1.00*** | 1.00 | 1.00 | 1.01** | 1.00 | 1.00 |

**Table S2: Regression outcomes for portal use before COVID-19 pandemic**

| **Outcome** | **Overall portal use** | **Messag-ing** | **Visits** | **My Record** | **Medical Tools** | **Billing** | **Proxy** | **Prefere-nces** | **Custom** |
| --- | --- | --- | --- | --- | --- | --- | --- | --- | --- |
| **Episode number** | 1.04 | 0.68*** | 1.16*** | 1.08*** | 0.93 | 1.08** | 1.15* | 0.95 | 1.10 |
| **Charlson**  **Co-morbidity index** | 1.02 | 1.08*** | 1.02 | 1.04** | 0.92** | 0.93** | 1.20** | 0.97 | 1.00 |
| **SDoH Ranking**  (Reference: Q1) |  |  |  |  |  |  |  |  |  |
| Q2 | 1.00 | 1.04 | 0.99 | 1.00 | 1.29** | 1.02 | 0.89 | 0.99 | 1.03 |
| Q3 | 0.97 | 1.01 | 0.97 | 0.97 | 1.10 | 0.94 | 1.02 | 0.90 | 0.96 |
| Q4 | 0.89** | 0.90** | 0.88** | 0.92** | 1.10 | 0.86*** | 0.79** | 0.88** | 0.92 |
| Q5 | 0.84*** | 0.91** | 0.88** | 0.88*** | 0.94 | 064*** | 0.95 | 0.95 | 0.96 |
| **Risk Category** (Reference=Normal) | 1.45*** | 1.32*** | 1.58*** | 1.48*** | 1.31*** | 1.30*** | 1.12 | 1.08 | 1.31** |
| **Race**  **(**Reference: Non-Hispanic White) |  |  |  |  |  |  |  |  |  |
| Non-Hispanic Black | 0.73*** | 0.66*** | 0.79*** | 0.76*** | 0.79** | 0.51*** | 0.93 | 1.08 | 0.97 |
| Hispanic | 1.06 | 0.86* | 1.20** | 1.00 | 1.23 | 1.00 | 0.61** | 1.29** | 1.41** |
| Others | 1.21 | 1.02 | 1.28* | 1.17*** | 1.18** | 1.34*** | 1.16 | 1.30*** | 1.40*** |
| **Age at first gestational encounter** | 1.00 | 1.01*** | 0.99*** | 1.00 | 1.01 | 1.01*** | 1.01 | 0.99** | 0.96*** |
| **BMI** | 1.00** | 1.00 | 1.01*** | 1.00** | 1.00 | 1.00 | 1.00 | 1.00 | 1.00 |
